# An Analysis of Oxycodone and Hydrocodone Distribution Trends in Delaware, Maryland, & Virginia Between 2006-2014

**DOI:** 10.1101/2022.07.16.22277708

**Authors:** Conor M. Eufemio, Joseph D. Hagedorn, Kenneth L. McCall, Brian J. Piper

## Abstract

**Objective:** Understanding opioid use and distribution trends by geographic area is critical in addressing the ongoing opioid epidemic in the United States. This study is a county level analysis of oxycodone and hydrocodone use in Delaware, Maryland, and Virginia between 2006-2014.

**Materials and Methods:** A retrospective analysis of oxycodone and hydrocodone distributed as collected by the Drug Enforcement Administration’s (DEA) Washington Post Automation of Reports and Consolidated Orders System (ARCOS) in Delaware, Maryland, and Virginia. Raw drug weights in each county were adjusted to “daily average dose” (grams/county population/365). Purchasing data collected from ARCOS was used to compare distribution trends during this period.

**Results:** There was a 57.59% in the weight of oxycodone and hydrocodone between 2006-2014. Oxycodone prescriptions increased by 75.50% and hydrocodone by 11.05%. Oxycodone increased across all three states between 2006-2010 and declined until 2014. Hydrocodone also increased but to a lesser extent than oxycodone. There was substantial variability in daily average dose of both opioids at the county level in all states. Pharmacies accounted for largest portion of oxycodone (69.17%) and hydrocodone (75.27%) purchased in the region. Hospitals accounted for 26.67% of oxycodone and 22.76% hydrocodone purchased. Practitioners and mid-level providers did not significantly contribute to this increase.

**Conclusion:** In the states of Maryland, Delaware, and Virginia, the distribution of the prescription opioids oxycodone and hydrocodone increased by 57.59%. Daily average dose increased between 2006-2010 in all three states followed by a decline until 2014. Variability in daily average dose by county highlights the relationship between geography and likelihood of receiving high dose opioids. It may further allude to effects of targeted distribution by pharmaceutical manufacturers and prescribing habits of geographically distinct healthcare entities. Relationships between location and opioid usage should continue to be investigated to promote rational use of controlled substances.

## Introduction

Oxycodone and hydrocodone are the number one and four opioids for annual mean consumption worldwide [1]. Along with other opioid derivatives, these drugs have been widely used in US inpatient and outpatient facilities for the treatment of both acute and chronic pain. The Food and Drug Administration (FDA) has approved them for moderate to severe pain and, as such, they are regularly prescribed for cancer-related, neurologic, and end-of-life pain [2].

Narcotics are often employed for pain management following surgical interventions. Approximately 51 million Americans undergo inpatient surgery annually. Four-fifths of prescription pain relievers involved either oxycodone or hydrocodone [3]. The rise of the US opioid epidemic has raised concerns about clinicians overprescribing habits and subsequent drug diversion to recreational use by patients. Regulations have sought to address these issues through improved accounting of prescription quantities and dosages administered to patients [4].

Opioids have been documented to increase dependence especially in the prolonged treatment of chronic pain [2]. The debilitating effects chronic pain have made opioids one of the most prescribed medication classes in the United States. In 2014, 245 million prescriptions for opioids were filled at retail pharmacies and 3-4% of adults were given long term opioid therapy. Diversion and improper use followed this increased availability. In 2013, more than 37% of the 44,000 overdose deaths were attributed to pharmaceutical opioids compared to 19% attributed to street grade heroin that year [5]. Post-operative pain management interventions are partially to blame for these events. Following procedures, prescriptions often go unused, are stored in unlocked spaces, and are not properly disposed creating a reservoir for misuse and subsequent injury and death [6].

Numerous studies have demonstrated variability in the prescription of controlled substances by geographic location [7-9]. Thus, specific analysis is warranted regarding prescribing trends in notable areas including the Mid-Atlantic states of Delaware, Maryland, and Virginia. Despite a similar location along the east coast, their respective sizes, population densities, and regulatory environments differ from one another. In 2018, Delaware attributed 88% of its 401 overdose deaths to opioids. Providers wrote 60.6 opioid prescriptions per 100 residents compared to the US average of 51.4. Maryland providers wrote 45.1 opioid prescriptions per 100 people and Virginia providers wrote 44.8 [10]

Understanding regional and national opioid prescribing trends is critical to our understanding of the ongoing opioid epidemic and may allow for improved utilization of public resources to resolve these issues. This report utilized the US DEA’s Automated Reports and Consolidated Ordering System (AROCS) comprehensive data base to evaluate changes in the use of oxycodone and hydrocodone in Delaware, Maryland, and Virginia between 2006-2014. Total weights of each medication were examined by county in each state. Regional analysis was undertaken using daily average dosage to investigate change between peaks years of drug usage. We also explored variations in the amount of these narcotics purchased by pharmacies, hospitals, practitioners, and mid-level providers as reported by ARCOS. This article was previously presented as a poster at the 2022 Pennsylvania Pain and Addiction Summit on April 19^th^, 2022.

## Material & Methods

Data was extracted from the DEA’s Washington Post ARCOS database which reports the monthly and quarterly weights of controlled substances prescribed along with purchasing data from distributors [11]. This included total grams of oxycodone and hydrocodone prescribed in Delaware, Maryland, and Virginia from 2006-2014. This resource has been frequently used in prior pharmacoepidemiologic studies [12-14]. The main units of this investigation may be less familiar than others such as “number of prescriptions.” Prior studies have indicated that examining the total weight of oxycodone in this data base relative to reports in a Prescription Drug Monitoring Program revealed a high correlation (R = .99) [12].

Our goal was to examine changes in opioid distribution and average daily doses by county in these states. However, ARCOS data is limited to raw weights distributed to providers and does not include statistics on the number of patients receiving opioid therapy or the average weight and duration of each prescription. To approximate the changes in each states relative use, the total weight of drug prescribed by county was divided by the county’s population (grams/population/365). The raw weights were adjusted to morphine equivalent weights (hydrocodone = 1.5 g*(reported weight) and oxycodone= 1 g*(reported weight)). Population statistics for each state, by county, were obtained from the US Census Bureau, however, inconsistencies were noted in the years of reporting [15]. Missing data was substituted with population statistics from the St. Louis Federal Reserve which revealed similar population trends to those already established [16]. This yielded a standardized dosing comparison on which further evaluation could be completed. These values were then used to determine changes in distributed weights from 2006-2010, a period where total weights of opioids increased throughout all states. Heat maps were generated using Someka® Excel software.

An examination of states specific buyer data was undertaking using reports from the DEA’s ARCOS. Reports included yearly purchased weight were used to compare the rates of distribution from different healthcare entities. This system tracks controlled substances distributed to pharmacies, hospitals, practitioners, and mid-level providers nationwide. Data was sorted for our states of interest. Institutional Review Board approval was by the University of New England and Geisinger. This article was previously posted to the medRxiv preprint server on July 26^th^, 2022.

## Results

Oxycodone use in Delaware, Maryland, and Virginia increased between 2006 and 2014 by 76.5% and hydrocodone use increased by 11.05%. Overall, this yielded a total increase of 57.59% (t=1.0394, p=0.3573, Figure 1). There was an increase in oxycodone distribution within the region between 2006-2010 (t=1.8317, p=0.1409). In Delaware, daily average dosages increased by +154.38% (t=2.50, p=0.0666). Daily dosages in Kent County accounted a +146.70% (t=1.9654, p=0.0670) change and Sussex County showed a significant +185.44% change (t=2.1461, p<0.05). New Castle also followed this trend with a +111.98% (t=5.5367, p<0.05) increase. Maryland’s statewide average daily oxycodone dose increased 84.51% between 2006 and 2010 (t=3.6593, p<0.05). Daily average doses continued increasing through 2011. There was substantial variability between counties as indicated by Figure 2. A pattern of higher per capita usage was noted around northern Baltimore suburban counties, and along both sides of the southern Chesapeake region. Somerset county presented the largest increase in daily average dose and a +213.45% increase between 2006 and 2010 (t=3.3156, p<0.05). High increases in usage were also observed in Cecil (+206.90%, t=3.1725, p<0.05), and Wicomico (+156.16%, t=2.0114, p=0.0791). These counties lie in the along the border with Delaware, with Wicomico and Somerset are located close to Sussex County. Virginia’s statewide daily average dosage of oxycodone increased by +52.96% (t=3.7637, p<0.05) between 2006 and 2010. However, the largest year over year increase came from the 2010-2011 reporting year (+18.15%). Several counties presented with increases greater than two standard deviations above Virginia’s state average, including Greene (+1272.50%) (t=7.4650, p<0.05), Grayson (+400.60%, t=9.4350, p<0.05), and Cumberland (+384.10%, t=12.6511, p<0.05). Like Maryland, Virginia showed substantial variability among counties as evidenced by the Figure 2. There do not appear to be significant correlates between geographic location and increase in daily average dose in Virginia. Many counties in northern Virginia increased in daily average dose above the state mean between 2006-2010, but not above a standard deviation. Counties with similar increases were seen in closer proximity to the West Virginia boarder or central Virginia as opposed to the coastal regions. It was noted that several of the counties reporting the largest increases daily average dose between 2006 and 2010 were located on the border with North Carolina (Figure 2).

**Figure 1:**
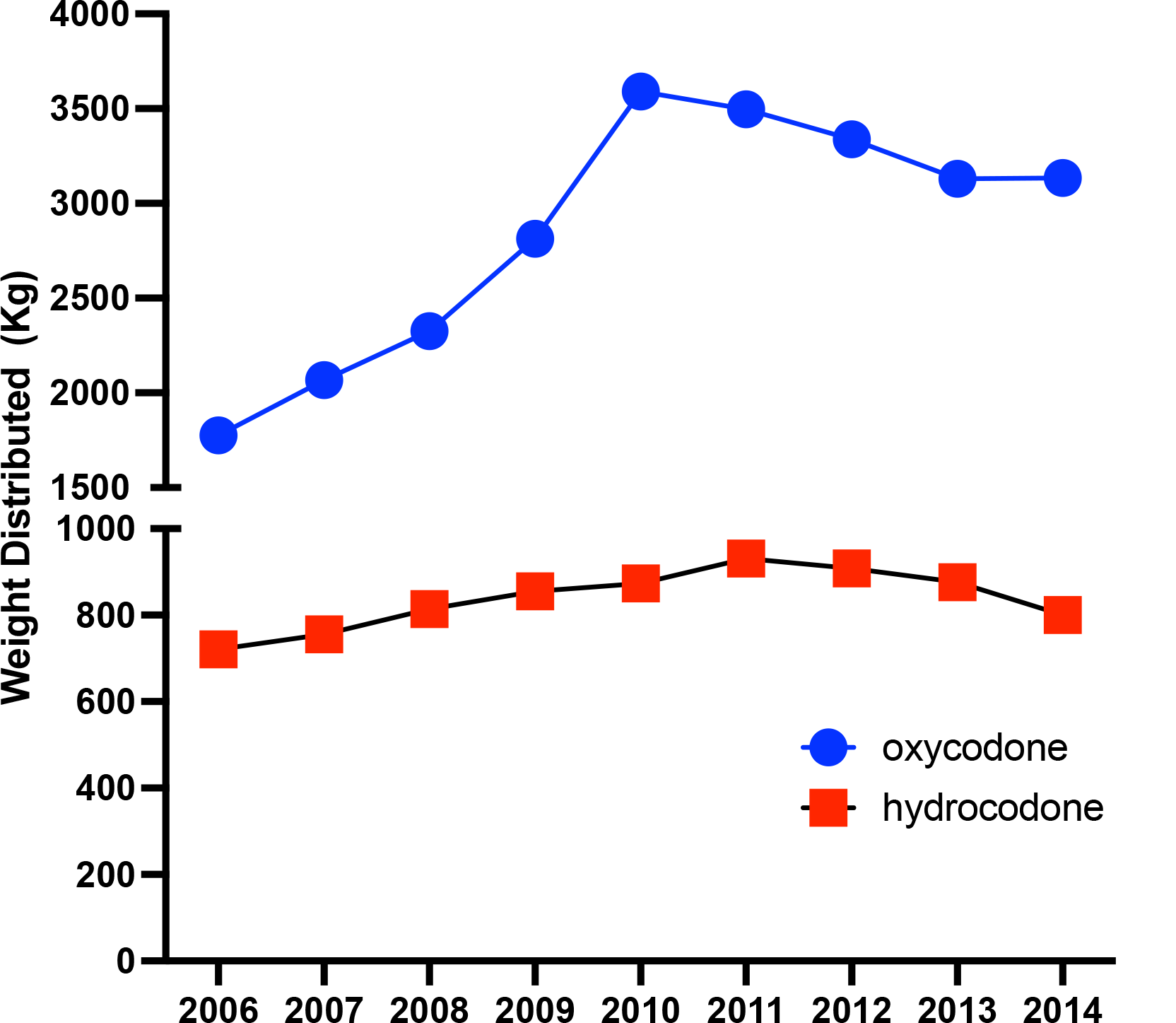
Trends in oxycodone and hydrocodone weights prescribed in Delaware, Maryland, and Virginia (2006-2010)

**Figure 2:**
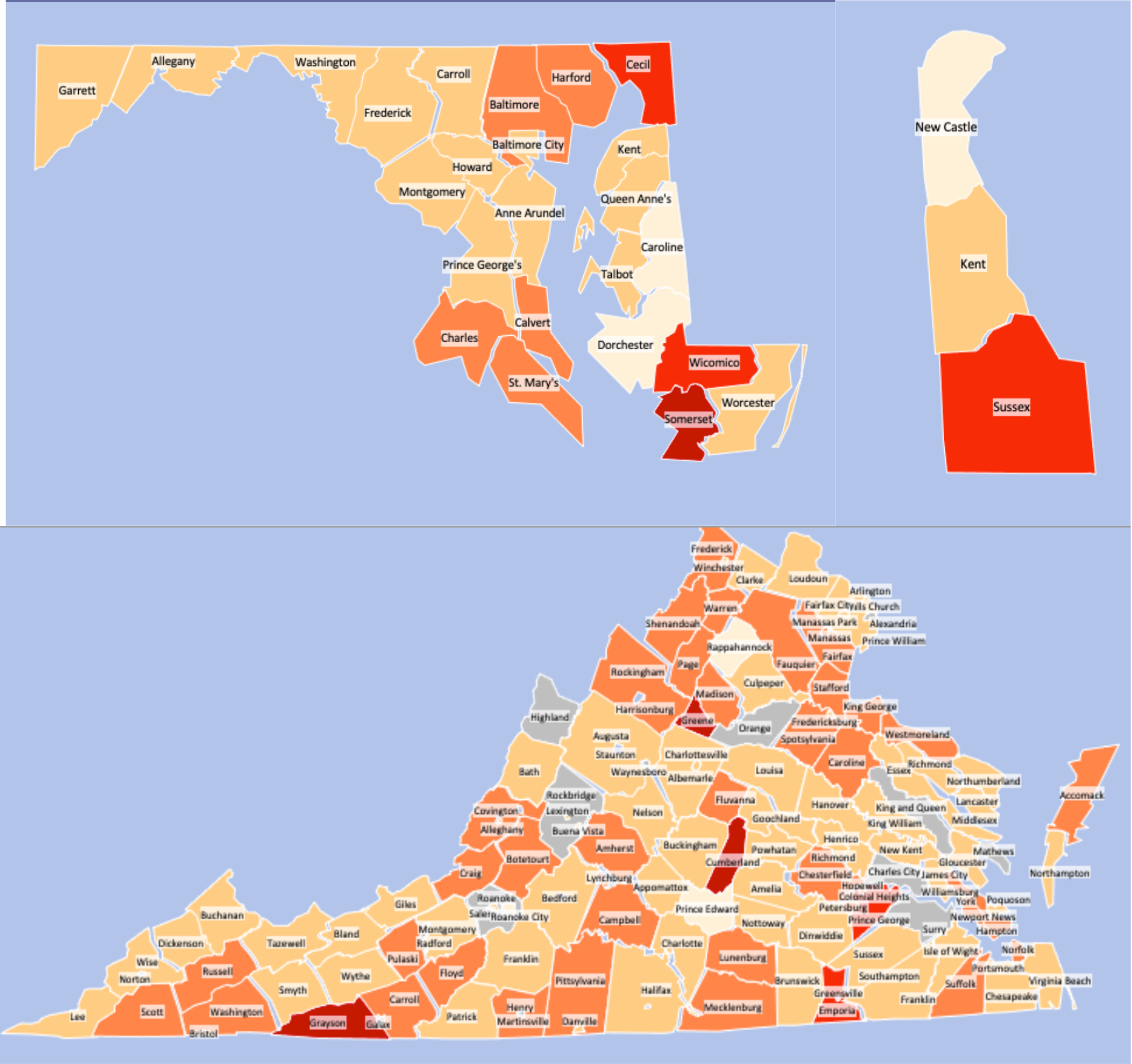
% change in oxycodone MME by county as reported by ARCOS (2006-2010) *Darker Red = More standard deviations above state mean *Grey = Population data not available

Trends in hydrocodone use remained steady across all three states. Delaware decreased its average daily dose by -14.91% between 2006 and 2014 (t=1.0162, p=0.3670). Both Kent (-20.06%; t=20.5349, p<0.05) and New Castle (-29.36%; t=21.6495, p<0.05) decreased their daily average dosage with only Sussex County (+4.69%; t=16.5075, p<0.05) showing an increase in distribution during this time. Maryland showed a slightly increased daily average hydrocodone dose (+4.82%) between 2006 and 2014. Most Maryland counties showed a decrease in daily average dose but there were significant increases in Garrett (+79.36%; t= 2.2892, p=0.05), Allegany (+33.18%; t=11.0918, p<0.05), Washington (+32.70%; t=4.6917, p=0.05), Frederick (+31.39%; t=9.9558, p<0.05) counties. The most significant reduction occurred in Baltimore City (-77.11%; t=7.2183, p<0.05). However, Virginia showed an overall increase of +36.52% in daily average dose between 2006 and 2014. Many of the contributing counties were in central and southern regions of the state and appear in clusters. There was significant variability between Virginia counties. When considering only the counties that increased their daily average hydrocodone dose, there was a 240-fold difference between the lowest (+2.36%) and highest (+568%) increase (Figure 3).

**Figure 3:**
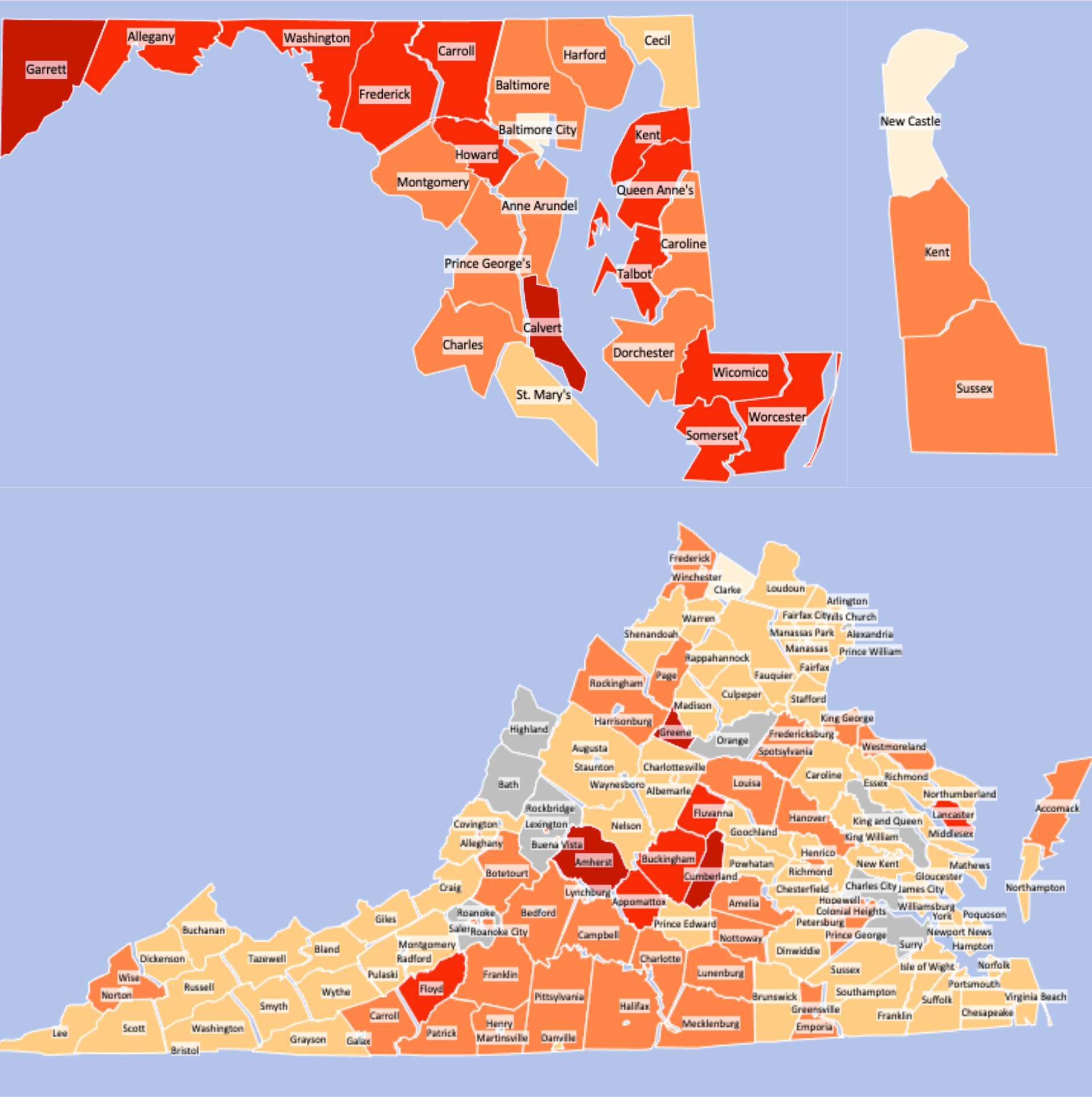
% change in hydrocodone MME by county as reported by ARCOS (2006-2010) *Darker Red = More standard deviations above state mean *Grey = Population data not available

**Figure 4:**
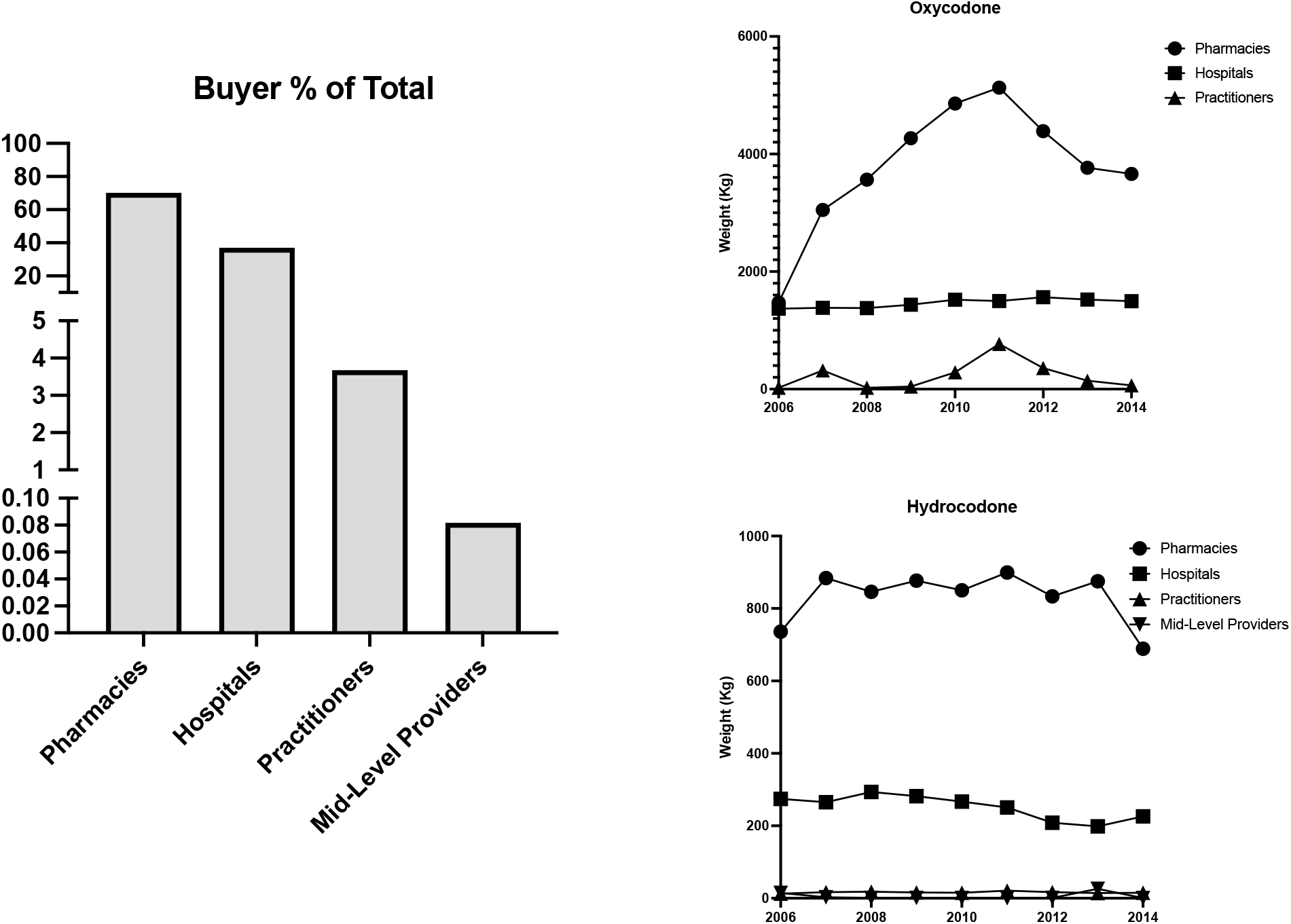
Buyer trends for oxycodone and hydrocodone in Delaware, Maryland, and Virginia.

ARCOS catalogues the weights of controlled substances bought by pharmacies, hospitals, practitioners, and mid-level providers. Pharmacies where responsible for purchasing over two-thirds of oxycodone (69.17%; t=3.1698, p<0.05) and three-quarters of hydrocodone (75.27%, t=7.9568, p<0.05). Between 2010-2011, pharmacies purchased +248.27% (t=7.3787, p<0.05) more oxycodone and +22.18% (t=12.5537, p<0.05) more hydrocodone. Hospitals followed in both oxycodone (+26.67%) and hydrocodone (+22.76%) purchasing. Practitioners increased the amount of oxycodone (+3,170.37%) and hydrocodone (+63.12%) between 2010-2011. However, however this portion of the total, 4.12% of oxycodone and 1.47% of hydrocodone, did not appreciably affect the cumulative trends. Mid-level providers only accounted for 0.49% of hydrocodone purchased between 2006-2014. Between 2010-2011, this decreased by -98.12%.

## Discussion

This study investigated trends in oxycodone and hydrocodone weights prescribed in the US states of Delaware, Maryland, and Virginia between 2006-2014, and indicated a 53.59% increase in the weight of oxycodone distributed. Analysis for total weight and daily average dose (adjusted for population) showed a comparable increase in oxycodone usage with a relatively small change (+11.05%) in hydrocodone usage.

Reduction in use following peak years may be reflective of new data that has come to light involving the relationship between opioid dose and risk of overdose [17]. Regulatory intervention likely influenced practitioners to adjust prescribing habits across the US. The peak year for opioid prescriptions occurred 2010-2011 in adjusted MME form. Additional data from the CDC reported that between 2006 and 2015, MME peaked across all opioid classes at 782 mg per capita followed by a gradual decrease which still left per capita prescribing weights three times greater than in 1999 [18].

Differences in distribution patterns related to geography has been the subject of research since the institution of prescription monitoring programs. Overall, studies have shown that counties located in the Appalachia region, Southern, and Western states are more susceptible to high numbers of prescriptions per person. These statistics are correlated with the larger county populations and proportions of residents who identify as non-Hispanic/African American or are poor, uninsured, and living in urban areas [7]. There have also been studies indicating that the primary driver for large quantities of opioid prescriptions is unrelated to patient characteristics such as the number of office visits or insurance coverage [19]. Other correlates including the percentage of residents living in poverty in 2010 were examined and compared to oxycodone weight prescribed between 2010 and 2011. 9.7% of Greene County, VA residents reported living below the poverty line at this time. An appreciable portion of residents in Mecklenburg (20.2%) and Southampton (16.4%) were living below the poverty line. The Virginia state poverty rate was 11.1% of residents. With two out of three of the highest prescribing counties above average poverty rates, this seems to be a more accurate indicator for likelihood of high daily average oxycodone dosage during this spike. The percentage of Delaware residents living in poverty was estimated to be 11.9% in 2010. Kent (12.80%) and Sussex (14.12%) were the two counties with above average poverty rates and the only counties where the weight of oxycodone prescribed had increased. However, New Castle’s reported poverty rate was a 11.2% and demonstrated a comparatively moderate increase of 4.40% in usage. Thus, in Delaware the poverty status of residents does not necessarily predict oxycodone dosage. The same is true for Maryland. Frederick, MD (5.6%) and Harford, MD (6.9%) both had poverty rates lower than 2010 state average (9.9%) despite reporting large increases in oxycodone dosage. Although poverty may be a factor the dosage of a prescription, this is likely confounded by other factors such as availability of primary care physicians, average ages of residents, and employment status may play a more important role in this correlation.

Additionally, we sought to explore changes in oxycodone distribution methods by evaluating ARCOS buyer data. ARCOS maintains yearly records on the weight of controlled substances purchased by pharmacies, hospitals, practitioners, mid-level providers, and teaching institutions across the US. Understanding opioid distribution patterns is key in designing protocol aimed at limiting demand in areas at high risk of overprescribing. Many states have implemented laws in recent years designed to strictly regulate the methods for prescribing opioids and other scheduled substances. In Delaware, the “Uniform Controlled Substances Act” was enacted in year and places clear definitions on administration and dispensing while also laying out titles for prescribers for which the law applies including physicians, dentists, and nurse practitioners. As defined by this law, all prescriptions, must be verified by a licensed provider regardless of circumstance. For example, emergent prescriptions must be verified by a board-certified physician [20].

Similar laws in other Mid-Atlantic states apply to prescribers and dispensers. Virginia does not allow for a narcotic supply exceeding seven days without clearly documented and affirmed medical records. However, this can be extended to 14 days following a surgical procedure. The state law also indicates specific measures to be taken with high doses or recurrent use.

Prescriptions above 120 MME require reasonable justification or a pain management referral. Naloxone is also prescribed to patients with risk factors including a prior overdose, substance misuse disorder, or concomitant benzodiazepine use [21]. In Maryland, doses >50 MME require an increase in follow up frequency and for naloxone to be offered to address overdose concerns. Pain management is to be consulted with doses >90 MME [25]. With emphasis on provider data, similar interest was taken in entities buying controlled substances. It should also be noted that prior ARCOS research has found that prescribing laws had no appreciable impact in Maryland and Delaware [23].

Within the region, pharmacies were the largest buyers of both oxycodone (69.2%) and hydrocodone (75.3%). This represented 70.2% opioid weight bought between 2006 and 2014. Hospitals were the second largest buyer of oxycodone (26.7%) and hydrocodone (22.8%). Year to year trends showed that oxycodone purchases by weight substantially increased between 2006 and 2011 between pharmacies in the state. In the peak year for average oxycodone dosage, 2011, pharmacies increased purchasing by +248.3% from 2006, compared to a +9.64% increase in hospital purchasing. This substantial increase likely associated with increased demand for outpatient opioid treatment. These trends fall in line with national analysis by the FDA which found an average increase of 15 billion MME per year supplied to retail pharmacies [24]. ARCOS data provided limited information on the type of pharmacy in question. This complicates trend analysis when evaluating changes in corporate vs independent pharmacy distribution patters. Delaware saw a +95% increase in the number of independent pharmacies operating in the state between 2010 (19) and 2019 (37) which was the largest increase in the United States during this period. Smaller increases were seen in Maryland (+36.0%) and Virginia (+8.0%) [25].

There was additional interest in purchasing trends of practitioners and mid-level providers. In total, practitioners in Delaware, Maryland, and Virginia, increased oxycodone purchase weight (+3170.4%) and hydrocodone weight (+63.1%) between 2010 and 2011. This describes 4.11% and 1.47% of purchase weights between the two medications, respectively. Consistent data for mid-level providers was only available for hydrocodone and represented a decrease of -98.1% during peak years accounting for <1% of hydrocodone purchased. Mid-Level providers, specifically Nurse Practitioners (NP) and Physician’s Assistants (PA), are increasingly autonomous in key areas of clinical care. In 2006, they accounted for 12.7% of emergency department patients seen, which was an increase from 5.5% in 1997 [26]. In all three states of interest, NPs and PAs are permitted to prescribe and dispense samples of Schedule II-V substances. NPs are allowed to procure in all three states while Virginia is the only one that allows NPs to do so. Other differences in restriction include those placed on ambulance services, chiropractors, and optometrists [27].

National studies have suggested that NPs/PAs prescribe opioids in similar patterns compared to MD’s, however, they were more likely to give high-frequency high-dose regimens [28]. Though procurement by mid-level providers was evaluated in this study, the possibility that mid-level providers are writing for opioid regimens in hospital settings cannot be discounted. As the US ranked third among all countries for highest per capita consumption of prescription opioids [1], further data to guide the policies of opioid stewardship programs are needed.

Thorough investigation is also needed to evaluate the relationship between diagnosis and opioid prescriptions particularly in rural areas that saw the highest population adjusted usage in these states. It was noted that counties with lower population densities saw the most significant increases in dosage by percent in their respective states. This finding may indicate the need for a more specified analysis of the clinical use of opioids in rural health centers to confirm other studies. For example, pervious analysis has suggested that there was an undertreatment of cancer-related pain in rural areas of Virginia in 2015 [29]. ARCOS data does not catalogue ICD-10 codes and therefore conclusions about prescriber behavior could not be drawn on the case of diagnostic trends. However, this information is imperative towards identifying the prescribing habits of providers and improving these trends in the interest of public health and patient safety. Other variables such as access to specialized oncologic care, diagnostic rates of terminal cancer, and local attitudes towards terminal pain treatment should also be considered.

Therefore, future studies, drawn from smaller cohorts, could analyze the relationship between geography, conditions, and use of opioids.

Overall, this analysis demonstrates showed three contiguous states with significant variability in the prescription of oxycodone and hydrocodone between their counties. These findings imply the need for federal, state, and local resources and policies to limit the use of these medications to address the opioid epidemic. This variability suggests that the need for resources from state and local funding for monitoring or treatment be handled by county authorities and health systems as opposed to regional ones. It stands to reason the that the maximum benefit from these interventions may be obtained by these channels

## Conclusions

In conclusion, this report identified increases in distribution of oxycodone and hydrocodone in Delaware, Maryland, and Virginia using ARCOS data. Examination revealed a combined 57.59% increase. Peak years for high dosage occurred between 2010-2011 which followed a substantial rise in daily average opioid dose in the region. Further investigation revealed that pharmacies were the largest suppliers of both substances, followed by hospitals. Practitioners and mid-level providers also increased the weight of opioids bought during this study period. Further state-level investigations are needed to better understand the economic and social factors influencing prescription trends and the effects of geography on substance overuse.

## Data Availability

All data present in the study are available upon request of the authors.
All data produced in the present work is contained in the manuscript.
All referenced data is from pubmed indexed sources

https://www.deadiversion.usdoj.gov/arcos/retail_drug_summary/index.html

## Contributor Roles

Conor M. Eufemio performed literature search and review, analyzed data, prepared figures, authored drafts of the paper, and approved the final manuscript.

Joseph D. Hagedorn performed literature search and review, retrieved and organized data for synthesis and analysis, and approved the final manuscript.

Kenneth L. McCall provided feedback on data analysis and interpretation and approved the final manuscript.

Brian J. Piper authored drafts of the paper, analyzed data, and approved the final manuscript.

## Institutional Review Board Statement

This study was deemed exempt by the IRB of Geisinger 2020-0223.

## Acknowledgements

This study was supported by the Health Resources Administration (D34HP31025).

